# Relative vaccine effectiveness (rVE) of mRNA COVID-19 boosters in people aged at least 75 years in the UK vaccination programme, during the Spring-Summer (monovalent vaccine) and Autumn-Winter 2022 (bivalent vaccine) booster campaigns: a prospective test negative case-control study

**DOI:** 10.1101/2023.03.16.23287360

**Authors:** Anastasia Chatzilena, Catherine Hyams, Rob Challen, Robin Marlow, Jade King, David Adegbite, Jane Kinney, Madeleine Clout, Nick Maskell, Jennifer Oliver, Adam Finn, Leon Danon, The Avon CAP Research Group

**Affiliations:** Engineering Mathematics, University of Bristol, Bristol, UK; Population Health Sciences, University of Bristol, Bristol, UK; Clinical Research and Imaging Centre, UHBW NHS Trust, Bristol, UK; Bristol Vaccine Centre, Population Health Sciences, University of Bristol, Bristol, UK; Academic Respiratory Unit, University of Bristol, Southmead Hospital, Bristol, UK; Bristol Vaccine Centre, Cellular and Molecular Medicine and Population Health Sciences, University of Bristol, Bristol, UK

**Keywords:** COVID-19, SARS-CoV-2, respiratory infection, vaccination

## Abstract

**Background:** Understanding the relative vaccine effectiveness (rVE) of new COVID-19 vaccine formulations against SARS-CoV-2 infection is an urgent public health priority. A precise analysis of the rVE of monovalent and bivalent boosters given during the 2022 Spring-Summer and Autumn-Winter campaigns, respectively, in a defined population has not been reported.

**Aim:** We therefore assessed rVE against hospitalisation for the Spring-Summer (fourth vs third monovalent mRNA vaccine doses) and Autumn-Winter (fifth BA.1/ancestral bivalent vs fourth monovalent mRNA vaccine dose) boosters.

**Methods:** A prospective single-centre test-negative design case-control study of ≥75 year-olds hospitalised with COVID-19 or other acute respiratory disease. We conducted regression analyses controlling for age, sex, socioeconomic status, patient comorbidities, community SARS-CoV-2 prevalence, vaccine brand and time between baseline dose and hospitalisation.

**Results:** 682 controls and 182 cases were included in the Spring-Summer booster analysis; 572 controls and 152 cases for the Autumn-Winter booster analysis. A monovalent mRNA COVID-19 vaccine as fourth dose showed rVE 46·6% (95% confidence interval [CI] 13·9-67·1) versus those not fully boosted. A bivalent mRNA COVID-19 vaccine as fifth dose had rVE 46·7% (95%CI 18-65·1), compared to a fourth monovalent mRNA COVID-19 vaccine dose.

**Conclusions:** Both fourth monovalent and fifth BA.1/ancestral mRNA bivalent COVID-19 vaccine doses demonstrated benefit as a booster in older adults. Bivalent mRNA boosters offer similar protection against hospitalisation with Omicron infection to monovalent mRNA boosters given earlier in the year. These findings support immunisation programmes in several European countries that advised the use of BA.1/ancestral bivalent booster doses.

## INTRODUCTION

Following the emergence of wild-type SARS-CoV-2 and circulation of antigenically distinct variants, large-scale vaccination programmes were implemented to reduce overall COVID-19 morbidity and mortality. In the UK, several COVID-19 vaccines received rapid regulatory authorisation: the vaccines used initially were the monovalent mRNA vaccine BNT162b2 (Cominarty®) and the ChAdOx1 (Vaxzevria®) replication-deficient simian adenovirus vector vaccine, with mRNA-1273 (Spikevax®) vaccine incorporated a few months later. These three COVID-19 vaccines were used in the primary campaign in the UK which began in December 2020, using initially an extended interval between first and second doses equal to 12 weeks, to prioritise first dose administration. The mRNA vaccines were offered as boosters 6-months after completion of the primary course, from September 2021for adults aged ≥50 years (y) and those in clinical risk groups, extending to all adults in November 2021 . A fourth dose of an mRNA vaccine was offered from March 2022 and prioritised the most vulnerable: all adults aged ≥75 y, and those in clinical risk groups[1]. A third priming vaccine dose had already been offered to immunosuppressed individuals, so that for them, the autumn 2021 and spring 2022 boosters were generally their fourth and fifth doses, respectively. The COV-Boost study indicated that a fourth-dose COVID-19 mRNA vaccination boosts immune responses[2], and an observational study showed three or four-dose vaccine effectiveness (VE) against hospitalisation of 60.9-62.1% against BA.4 or BA.5 variants which emerged during spring 2022 and 50.1% against BA.2 when compared to two-doses received ≥25 weeks earlier[3]. These initial COVID-19 vaccines were developed against wild-type virus and provided significant protection against infection, hospitalisation, severe disease and death[4–8]. However, VE has been eroded progressively both by waning of immune-protection over time and emergence of SARS-CoV-2 variants of concern (VOC) (Alpha, Delta, Omicron) which show immune escape, [9–12].

In summer 2022, the UK Medicines & Healthcare products Regulatory Agency (MHRA) approved two new bivalent booster vaccines which were developed in response to concerns about such viral evolution and escape. The Moderna bivalent vaccine (Spikevax® bivalent Original/Omicron) was approved on 15th August 2022, followed quickly by Pfizer/BioNTech bivalent vaccine (Comirnaty® Original/Omicron BA.1) approval on 3rd September 2022[13,14] and they were distributed during autumn 2022, being the fifth dose offered in the UK. The Moderna vaccine contains 25mcg of mRNA coding for the spike protein of the ancestral strain and 25mcg of mRNA against Omicron (BA.1) and the Pfizer BioNTech vaccine contains 15mcg of mRNA directed against the ancestral strain and 15mcg of mRNA against Omicron (BA.1). Early immunogenicity studies suggest bivalent mRNA boosters induce similar or higher neutralising antibody levels against Omicron sub-variants and other VOCs compared to monovalent mRNA boosters[15–19].

SARS-CoV-2 infection incidence remains high[20], whilst determining whether patients who test SARS-CoV-2 positive have COVID-19 has become increasingly challenging using studies relying on data linkage methodology. Additionally, comparison between vaccinated individuals and those who have not received any COVID-19 vaccine dose cannot be performed, as 78.2% of the adult population in the UK have received at least 2 doses or had prior exposure to SARS-CoV-2: thus even unvaccinated individuals have some immunity to SARS-CoV-2. There remains limited evidence[10,15,21] of bivalent vaccines’ clinical effectiveness when compared to monovalent formulations because the different vaccine rollout timings make a direct comparison of the vaccines impossible. Acknowledging this constraint, we undertook a test-negative design (TND) case control study comparing SARS-CoV-2 positive and negative acute lower respiratory tract disease (aLRTD) patients, implementing two separate analyses to assess the protection against SARS-CoV-2 hospitalisation provided by an additional monovalent or BA.1/ancestral bivalent mRNA vaccine dose relative to those who had not received the respective doses, focusing on ≥75y olds who were the main target group in the spring 2022 booster programme. Given the different rollout timings of the two vaccine formulations, the analyses refer to two distinct study time periods with different subvariants circulating.

## METHODS

### Study design and conduct

We estimated the relative vaccine effectiveness (rVE) of monovalent and bivalent mRNA vaccines against hospitalisation in Bristol, within the study population consisting of adults admitted to North Bristol and University Hospitals Bristol and Weston NHS Trusts [AvonCAP: ISRCTN17354061] between 4th April 2022 and 30th July 2022 (the period following the initiation of distribution of fourth dose of monovalent mRNA vaccines), and between 21st September 2022 and 23rd January 2023 (the period following the initiation of distribution of bivalent mRNA vaccines) inclusive. During the first study period BA.4, BA.2 and BA.5 were the main Omicron sub-lineages identified in COVID-19 cases in England, while the dominant sub-lineages identified during the second study period were BA.5, BA.4.6, BQ.1, CH1.1, XBB recombinant lineage and its mutation XBB.1.5 (Supplementary Data S2) [22]. The study population consisted of patients with signs/symptoms of respiratory infection, aged ≥18y at the time of hospitalisation[23]. Study eligible cases and controls eligible were identified from the medical admission list, and data were collected from medical records using REDCap software[24]. All data collection was undertaken by individuals not involved in analysis and blinded to results, following the same procedures for both cases and controls. Vaccination records for every study participant were obtained from linked hospital and GP records, including vaccine brand and date of administration with data collection performed by individuals blinded to participants’ SARS-CoV-2 test results[25].

### Case definition and exclusions

We included patients with ≥2 signs of acute respiratory disease (cough, fever, dyspnoea, tachypnoea, increased/discoloured sputum expectoration, pleurisy, clinical or radiological findings suggestive of acute disease) or a confirmed clinical/radiological diagnosis of aLRTD[25]. Patients hospitalised with aLRTD and positive admission SARS-CoV-2 test using the UK Health Security Agency (UKHSA) diagnostic assay in use at the time were classified as cases; those with aLRTD and negative SARS-CoV-2 result were classified as controls. Eligible controls could have multiple hospitalisations, provided subsequent admissions were >7 days following previous discharge. We included only the first COVID-19 admission for each case.

We excluded patients whose admission date was >10 days after symptom onset date (to avoid including potentially false negative admission SARS-CoV-2 tests), and those with a confirmed previous SARS-CoV-2 infection based on any positive test result that could be found in local and/or national clinical care record database, including linkage through the UKHSA national testing system. Patients who had received 2 vaccine doses or fewer at the time of admission were also excluded (Supplementary Data S1).

In order to make a side-by-side evaluation of the effectiveness of the different booster vaccine formulations, we restricted both analyses to individuals aged ≥75y, since the Joint Committee on Vaccination and Immunisation (JCVI) advised targeting COVID-19 booster vaccines during spring-summer towards those at highest risk of severe disease; those aged ≥75y and residents in long-term care facilities (LTCFs)[1], while in Autumn-Winter 2022 the offer was extended, including those aged ≥50y and frontline health and social care workers [26]. Therefore, patients < 75y were excluded from analyses.

### Exposure definition

This analysis aims to measure the protection offered by an additional dose of monovalent (Original ‘wild-type’ mRNA vaccine, Pfizer-BioNTech [Comirnaty®] or Moderna [Spikevax®]) and bivalent (Original ‘wild-type’/Omicron BA.1 mRNA vaccine, Pfizer-BioNTech [Comirnaty®] or Moderna [Spikevax®]) vaccine within 3 months after vaccination, each compared with those who had not received the respective boosters, side by side, during SARS-CoV-2 Omicron variant dominance. The Spring-Summer monovalent booster analysis (admissions 04/04/22-30/07/22) compares the fourth dose of monovalent given as a booster (21/03/22-07/08/22) in the UK, to the third dose of monovalent vaccine during Autumn-Winter 2021 (16/09/21-14/02/22). By the end of this study period, the vaccine uptake in the UK in the over 75s was 74.3% for spring 2022 booster and 93.5% for 3 doses (cf. median vaccine uptake of 13.6% and 84.2% respectively in EU/EEA countries in over 60 year olds, based on available data [27]). The Autumn-Winter bivalent booster analysis (admissions 21/09/22-23/01/23) compares the fifth dose of vaccine, with the bivalent formulation given as a booster (07/09/22-12/02/23) to the fourth dose of monovalent vaccine in Spring-Summer 2022 (21/03/22-07/08/22). The vaccine uptake in individuals aged ≥75y by the end of this study period was 83.5% for autumn 2022 booster and 74.9% for spring 2022 booster (cf. median vaccine uptake of 2.2% and 35.1% respectively in EU/EEA countries in over 60 year olds based on available data [27]).

For the Spring-Summer monovalent booster analysis, individuals were defined as boosted with a monovalent vaccine if they had received three doses of any monovalent vaccine combination and a fourth dose of monovalent vaccine during the Spring-Summer 2022 vaccination programme, and no more than three months prior to their admission, and as not fully boosted only if they had received two doses of any vaccine combination followed by a third dose of monovalent vaccine during Autumn-Winter 2021. For the Autumn-Winter bivalent booster analysis, individuals were defined as boosted with a BA.1/ancestral bivalent vaccine if they had received four doses of any vaccine combination plus a fifth bivalent dose during Spring-Summer 2022 vaccination programme and no more than three months prior to their admission, and as not fully boosted if they had received three doses of any vaccine combination plus a fourth monovalent dose during Autumn-Winter 2022. In both analyses, we define those having received the most recent dose with >7 days having elapsed between the vaccine and symptom onset as immunised (Supplementary Data S1).

Individuals who received a third vaccine dose in Autumn-Winter 2021, a fourth dose in Spring-Summer 2022 and a fifth dose in Autumn-Winter 2022, are those who had received two doses as the primary vaccination regimen before and during Spring-Summer 2021. However, individuals with severe immunosuppression around the time of their first or second vaccine doses were offered an additional primary dose (third dose) before any booster doses. As a result, they were offered a fourth vaccine dose in Autumn-Winter 2021, a fifth dose in Spring-Summer 2022 and a sixth dose in Autumn-Winter 2022 (Figure 1, Supplementary Data S2). Since this population almost exclusively comprised of immunosuppressed individuals we perform additional sensitivity analyses including those individuals who had received three doses as their primary vaccination regimen in both comparisons.

**Figure One:**
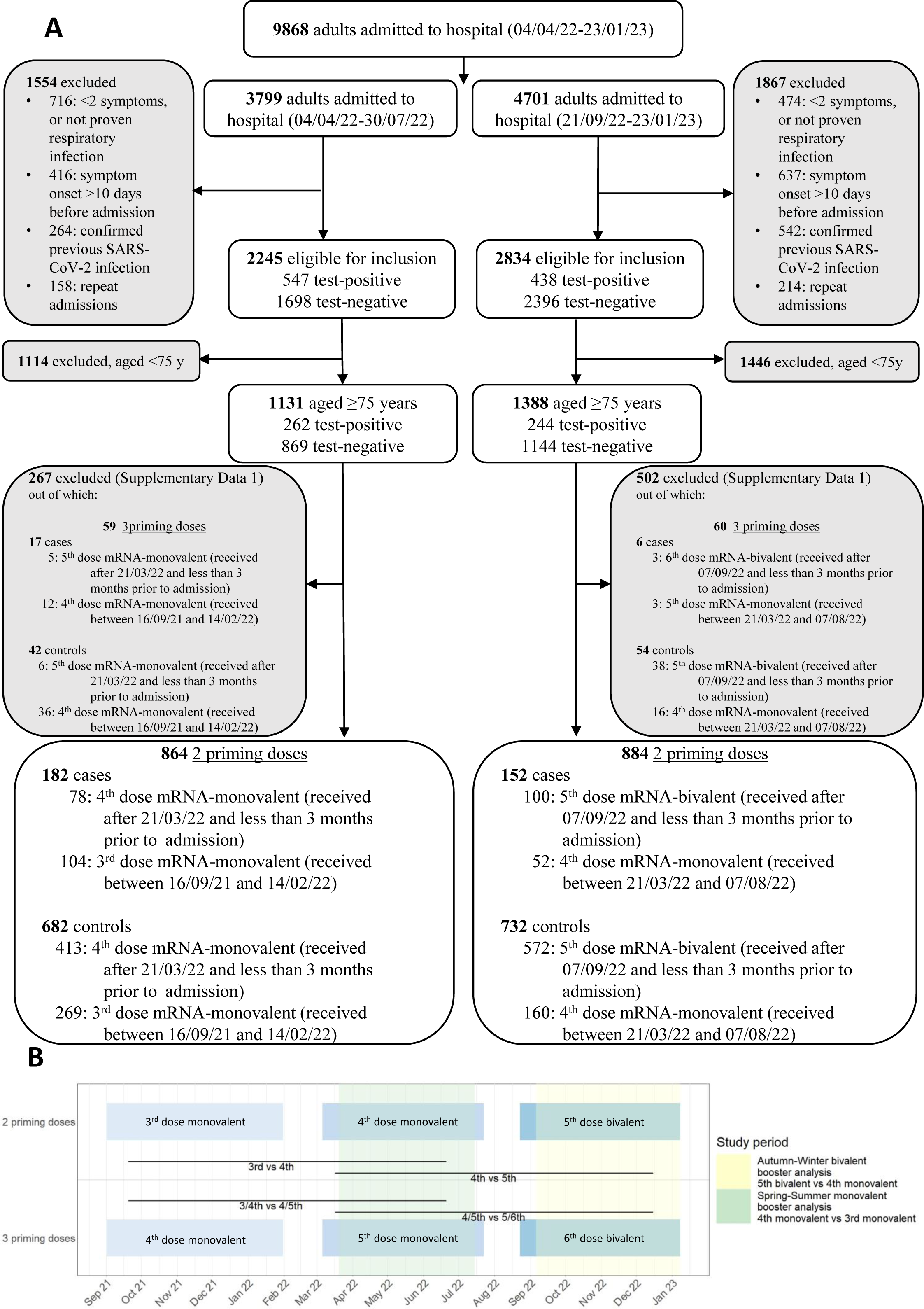
Study Flow Diagram. (A) Inclusion and exclusion criteria in the cohort. The left panel describes the Spring-Summer monovalent booster analysis; of the 182 SARS-CoV-2 positive individuals, 78 were vaccinated with 4th dose monovalent mRNA vaccine, and 104 were vaccinated with 3rd dose monovalent mRNA vaccine. Of the 682 SARS-CoV-2 negative individuals admitted, 413 were vaccinated with 4th dose monovalent mRNA vaccine and 269 were vaccinated with 3rd dose monovalent mRNA vaccine. The right panel describes the Autumn-Winter bivalent booster analysis; of the 152 SARS-CoV-2 positive individuals, 100 were vaccinated with 5th dose bivalent mRNA vaccine, and 52 were vaccinated with 4th dose monovalent mRNA vaccine. Of the 732 SARS-CoV-2 negative individuals admitted, 572 were vaccinated with 5th dose bivalent mRNA vaccine and 160 were vaccinated with 4th dose monovalent mRNA vaccine. (B) Study timeline

### Outcomes

We assessed the additional protection provided by a fourth dose of mRNA monovalent vaccine and a fifth dose of BA.1/ancestral bivalent vaccine as boosters against the primary endpoint of hospital admission with a positive admission SARS-CoV-2 test and either a clinical or radiological aLRTD diagnosis or aLRTD signs/symptoms compared to that provided by three or four doses of the monovalent formulations of the vaccines, respectively.

### Statistical Analysis

Demographic, clinical characteristics and other factors that may affect the exposure (vaccination status) or outcome (hospital admission) were compared between cases and controls for both comparisons, between monovalent vaccine boosted and not fully boosted, and between boosted with a bivalent vaccine and not fully boosted, using Fisher exact tests (categorical variables), two-sided Kolmogorov-Smirnov tests (continuous variables) and Wilcoxon rank-sum tests (score variables).

Under TND assumptions, we estimate the odds ratio of testing SARS-CoV-2 positive among patients boosted with a monovalent vaccine versus those not fully boosted (rOR) and define rVE as (1–rOR)×100. Similarly, we estimated rVE of bivalent booster, comparing the odds of testing positive for SARS-CoV-2 among patients boosted with a bivalent vaccine versus those not fully boosted. This was done using univariable logistic regression (Univariable Logistic Regression Model). Differences in the timing of third/fourth dose and rollout timings of different vaccine brands could introduce unobserved biases, confounding results in both comparisons. To mitigate these, we performed multivariable logistic regression analyses adjusting for time between baseline vaccine dose (third dose for the Spring-Summer monovalent booster, and fourth dose for the Autumn-Winter bivalent booster analysis) and admission (in days), vaccine brand [binary variable], age, sex, Index of Multiple Deprivations (IMD) decile rank and Charlson comorbidity index (CCI) [continuous variable], LTCF residency status, presence of pre-existing respiratory disease, and community SARS-CoV-2 prevalence lagged by time interval between infection and hospitalisation, assumed to be 8-days (Multivariable Logistic Regression Model).

We also conducted sensitivity analyses, matching cases and controls using propensity score balancing using logistic regression to define propensity score, and nearest neighbour matching. Matching variables included age, sex, CCI and IMD, LTCF residency status and presence of pre-existing respiratory disease, and likelihood of vaccine receipt (Matched Conditional Logistic Regression Model). Matching by elapsed time since baseline vaccine dose/brand was not performed to avoid introducing bias[28]. Time since baseline vaccine dose/ brand is affected by dose of last vaccine received; each booster was deployed ≥4 months after the previous COVID-19 booster programme (Figure 1), with different programmes using different proportions of each vaccine brand. As additional sensitivity analysis, we included individuals who had received three doses as primary vaccination regimen, adjusting for the number of primary doses [binary variable], using the same methods for both comparisons.

Statistical analyses were performed with R v4.0.2. Missing data were limited to the IMD variable and accounted for <1%; no imputation was performed; all analyses only included participants with complete data. Statistical significance was defined using a 2-sided significance level of α=0·05.

### Ethics and permissions

The Health Research Authority Research Ethics Committee (East of England, Essex), REC20/EE/0157 approved this study, including using Section 251 of the 2006 NHS Act under Confidentiality Advisory Group authorisation.

## RESULTS

During the periods evaluated, 9,868 adult aLRTD hospitalisations occurred in Bristol, UK, while the Omicron variant was dominant[22,29,30]. In the Spring-Summer booster, 864 admissions of ≥75y old patients hospitalised with SARS-CoV-2 aLRTD were eligible for this analysis: median patient age was 85y (IQR 80-89), 403 individuals (46%) were male, median CCI was 5 (IQR 4-6) with no significant differences in patient age and sex between SARS-CoV-2 positive and negative aLRTD, while there was a statistically significant difference in their ethnicity, LTCF residency status, smoking and presence of pre-existing respiratory disease and chronic obstructive pulmonary disease (Figure 1, Table 1). In the Autumn-Winter booster, 884 admissions of ≥75y old patients were eligible with no significant differences in age, sex and LTCF residency status between SARS-CoV-2 positive and negative aLRTD, while differences in ethnicity, smoking and dementia status were statistically significant (Figure 1, Table 2). In both comparisons, there were no significant differences in patient demographics and health status between vaccination groups (Supplementary Data S3, S4).

In the Spring-Summer booster analysis, of the 182 SARS-CoV-2 cases, 78 (43%) received a fourth monovalent vaccine dose and 104 (57%) received a third monovalent vaccine dose, while 413 of 682 controls (61%) received a fourth monovalent vaccine dose and 269 (39%) received a third monovalent vaccine dose. All those vaccinated with a fourth monovalent dose were hospitalised ≤3 months after their vaccination and 98% of those who had received only 3 doses were hospitalised >3 months after their vaccination (Supplementary Data S3). The unadjusted rVE was 51·2% (95%CI 32·1-65) and after adjustment, rVE was 46·6% (95%CI 13·9-67·1).

Matched conditional logistic regression rVE was 52% (95%CI 20·9-70·9) (Table 3). Sensitivity analysis including individuals with three doses as primary vaccination regimen (who make up 8.5% of cases and 5.8% of controls) resulted in lower rVE estimates compared with results from the main analysis (Table 3, Supplementary Data S5, S6, S9).

In the Autumn-Winter booster analysis, of the 152 SARS-CoV-2 cases, 100 (66%) received a fifth BA.1/ancestral bivalent vaccine dose and 52 (34%) received a fourth monovalent vaccine dose, while 572 of 732 (78%) controls received a fifth bivalent vaccine dose and 160 (22%) received a fourth monovalent vaccine dose. All those vaccinated with a fifth BA.1/ancestral bivalent vaccine dose were hospitalised ≤3 months after their vaccination and 97% of those who had received only 3 doses were hospitalised >3 months after their vaccination (Supplementary Data S4). The unadjusted rVE was 46·2% (95%CI 21·1-63) and after adjustment, rVE was 46·7% (95%CI 18-65·1). Matched conditional logistic regression rVE was 48·8% (95%CI 19·8-67·3) (Table 4). The inclusion of individuals with three doses as primary vaccination regimen (who make up 3.8% of cases and 6.9% of controls) produced estimates comparable with results from the main analysis (Table 4, Supplementary Data S7, S8, S10).

## DISCUSSION

In this analysis, we consider the public health implications of monovalent and BA.1/ancestral bivalent vaccine implementation, focusing on people aged ≥75y; a high-risk group which was a primary target for the UK vaccination programme. Although COVID-19 vaccines have been shown to be effective against severe COVID-19 disease[4,31,32], it has not been possible to compare the effectiveness of monovalent boosters directly with BA.1/ancestral bivalent booster doses of mRNA COVID-19 vaccines in defined populations, because bivalent formulations rapidly and entirely replaced monovalent formulations in the most recent booster programmes. In this ongoing prospective study, we undertake a sequential sub-analysis of the two vaccines given as boosters during two booster programmes in the same calendar year, providing evidence that monovalent vaccine (Original ‘wild-type’ Comirnaty® and Spikevax®) and BA.1/ancestral mRNA bivalent vaccine (Original ‘wild-type’/Omicron BA.1 Cominarty® and SpikeVax®) within three months after vaccination provided similar additional protection compared to that afforded by waned previous doses against hospitalisation from Omicron SARS-CoV-2 sub-variants in older individuals, given unknown effects of the different subvariant circulating during the distribution of the two vaccine formulations (Supplementary Data 2 C).

Specifically, we estimated that a fourth monovalent mRNA vaccine dose within three months after vaccination, was associated with a 46·6% (95%CI 13·9-67·1) additional protection against hospitalisation compared to waned three doses, in individuals ≥75y, during Omicron BA.2, BA.4, BA.5 lineage dominance. Within three months of receiving a fifth bivalent mRNA vaccine dose, it is estimated to provide 46·7% (95%CI 18-65·1) additional protection against hospitalisation compared to waned four doses, in the same ≥75y age group, even when assessed during a period when heterologous variants were circulating since BA.5, BA.4.6, BQ.1 and CH1.1 lineages, the XBB recombinant lineage and its mutation XBB.1.5 accounted for most of the identified cases in England (Supplementary Data 2 C) [22].

Although our results demonstrate that both vaccine formulations combined in these booster programmes had benefits when used as boosters, we have insufficient case numbers to draw conclusions about individual vaccine brands or directly compare them. Importantly, this analysis is restricted to individuals ≥75y old, with 97-98% of not fully boosted individuals in our sample potentially having waned vaccine-induced immunity since they received their last dose more than 3 months prior to admission(Supplementary Data S2, S3). Given that this study has a short follow up period after the administration of the two boosters, we cannot provide estimates by time since vaccination; nonetheless it presents encouraging evidence of similar benefit of monovalent and bivalent boosters in older adults, up to three months after vaccination. Older adults are at increased risk of severe disease, and protection may wane faster[33]; older adults were therefore targeted in the UK Spring-Summer 2022 and Autumn-Winter 2022 COVID-19 booster programmes. Our analysis based on the inclusion of individuals with severely weakened immune systems who were eligible for three primary doses in our basic comparisons, suggests that both vaccine formulations may offer some additional protection, however given that they account for <7% of our sample we have insufficient statistical power to draw firm conclusions. Vaccination against SARS-CoV-2 is independently associated with lower COVID-19 severity[4,31,32], and vaccines have been an important disease modifier during the pandemic. Our estimates suggest that the bivalent boosters provided similar protection as monovalent boosters in a real-world setting where the landscape of COVID-19 variants is constantly changing: results concordant with early evidence suggesting neutralising antibody titres induced against Omicron by a bivalent booster dose were not higher than following a monovalent booster dose in small studies[16,17]. Our results are concordant with a recent UKHSA analysis[34] which estimated the incremental protection conferred by a fourth monovalent dose compared to waned third dose was 58.8% (95%CI 54-63%), while the additional protection of BA.1/ancestral mRNA bivalent vaccines relative to those with ≥2 doses and waned protection was 57% (95%CI 48-65%), during the same time period as our analysis.

Since the study took place over the course of two different time periods, the interpretation of these sequential analyses of the two vaccine formulations has to take into account the different variants circulating[22,29,30]. In England, the Omicron variants BA.2, BA.4, BA.5 were the main circulating variants during the study period of the monovalent booster, and were replaced by BA.4 and BA.5 descendent sub-lineages(BA.4.6, BQ.1), CH1.1, XBB and XBB.1.5 lineage during the study period of the BA.1/ancestral bivalent boosters, with BA.5 being the only subvariant in common. Consequently, in this study, the performance of the BA.1/ancestral bivalent booster was not evaluated against homologous subvariants but against BA.4/5 which show further immune escape beyond that observed for BA.1. Currently, there is no evidence that Omicron BA.4-related sublineages, Omicron BA.5-related sublineages, CH1.1 and XBB recombinant-related sublineages, which appeared during the study period of the BA.1/ancestral mRNA bivalent boosters, cause more severe disease. The impact of these lineages on the effectiveness of the BA.1/ancestral mRNA bivalent formulation has not yet been studied in detail.

The TND has been described previously, along with its advantages and limitations[12,25,35], and our analysis has some important additional strengths and limitations. The strength of our approach is the focus on using real-world data, while accounting for the potential effects of LTCF residency status, socioeconomic status and comorbidities. By limiting our analysis to boosted individuals only, our analysis sidesteps the potentially unfair comparisons between populations that have followed UK COVID-19 vaccine recommendations and unvaccinated populations that may display other idiosyncratic behaviours. We also utilised symptom onset date to define illness start time and are able to confirm that there is no difference in time since vaccination between the case and control groups compared. We, therefore, define illness onset relative to both vaccination and hospitalisation date accurately, without relying on positive test date (which may vary widely), eliminating this source of bias or misclassification. All patients were hospitalised with acute respiratory illness, so these results are unlikely to be subject to significant bias caused by admission for other causes (i.e., incidental COVID-19 disease). Most notably, our estimates for the effect of individual vaccines are underpowered, due to small patient numbers in our cohort during the phases of the UK vaccination programme and study periods. We are unable to assess additional protection against other markers of disease severity, such as admission to intensive care or requirement for respiratory support, due to the small number of eligible admissions in this time period. This analysis does not measure rVE in individuals who were not hospitalised or were asymptomatic, so we cannot determine protection against asymptomatic disease or transmission. Treatment biases may result in community-based treatment, death before admission, or patients may otherwise not be referred to hospital. We note that this cohort, whilst broadly representative of the UK population, was predominantly Caucasian and the studied vaccines may have different effectiveness in individuals from other ethnic backgrounds.

In this prospective study, we provide evidence that autumn BA.1/ancestral mRNA bivalent COVID-19 boosters offered similar augmentation of protection against Omicron hospitalisation to that induced by spring monovalent mRNA boosters in 2022. These findings support immunisation programmes in the UK and several European countries, that advised the use of BA.1/ancestral mRNA bivalent booster doses in high risk individuals.

## Data Sharing

The data used in this study are sensitive and cannot be made publicly available without breaching patient confidentiality rules. The data dictionary is therefore unavailable.

