## Supplementary Data for "Relative vaccine effectiveness (rVE) of mRNA COVID-19 boosters in people aged at least 75 years in the UK vaccination programme, during the Spring-Summer (monovalent vaccine) and Autumn-Winter 2022 (bivalent vaccine) booster campaigns: a prospective test negative case-control study"

**Supplementary Data 1 (S1): Study design, inclusion and exclusion criteria**

|  | **Spring-Summer monovalent booster analysis [4^th^ dose – 3^rd^ dose monovalent mRNA vaccines] admitted to hospital (04/04/22-30/07/22)** | **Autumn-Winter bivalent booster analysis [5^th^ dose BA.1/ancestral mRNA bivalent - 4^th^ dose monovalent vaccines] admitted to hospital (21/09/22-23/01/23)** |
| --- | --- | --- |
| **Inclusion criteria** | - 4^th^ dose mRNA-monovalent received after 21/03/22 and less than 3 months prior to admission - 3^rd^ dose mRNA-monovalent received between 16/09/21 and 14/02/22 | - 5th dose mRNA-bivalent received after 07/09/22 and less than 3 months prior to admission - 4th dose mRNA-monovalent received between 21/03/22 and 07/08/22 |
| **Exclusion criteria** | - <2 symptoms, or not proven respiratory infection - symptom onset >10 days before admission - confirmed previous SARS-CoV-2 infection - repeat admissions | - <2 symptoms, or not proven respiratory infection - symptom onset >10 days before admission - confirmed previous SARS-CoV-2 infection - repeat admissions |
|  | - <75 years old | - <75 years old |
|  | - 4^th^ dose mRNA-monovalent received before 21/03/22 (4th dose during Autumn-Winter 2021 booster programme i.e. they had 3 primary doses) - 4^th^ dose mRNA-monovalent received after 21/03/22 and more than 3 months prior to admission - 4^th^ dose ChAdOx1 (Vaxzevria®) - 3^rd^ dose mRNA-monovalent received before 15/09/21 or after 15/02/22 - 3^rd^ dose ChAdOx1 (Vaxzevria®) - Vaccinated with 5 (5th dose during Spring-Summer 2022 booster programme i.e. they had 3 primary doses), 2 doses or 1 dose - Unvaccinated | - 5th dose mRNA-monovalent (5th dose during Spring-Summer 2022 booster programme i.e. they had 3 primary doses)5th dose mRNA-bivalent received after 07/09/22 and more than 3 months prior to admission - 5^th^ dose ChAdOx1 (Vaxzevria®) - 4^th^ dose mRNA-monovalent received before 20/03/22 or after 08/08/22 - 4^th^ dose ChAdOx1 (Vaxzevria®) - Vaccinated with 6 (6th dose during Autumn-Winter 2022 booster programme i.e. they had 3 primary doses), 2 doses or 1 dose - Unvaccinated |

**Supplementary Data 2 (S2): Admissions by vaccination status in each cohort by admission date – Admissions by vaccination status in each cohort by vaccination date**

| **Spring-Summer monovalent booster analysis [4^th^ dose – 3^rd^ dose monovalent mRNA vaccines] admitted to hospital (04/04/22-30/07/22)** | **Autumn-Winter bivalent booster analysis [5^th^ dose BA.1/ancestral mRNA bivalent - 4^th^ dose monovalent vaccines] admitted to hospital (21/09/22-23/01/23)** |
| --- | --- |

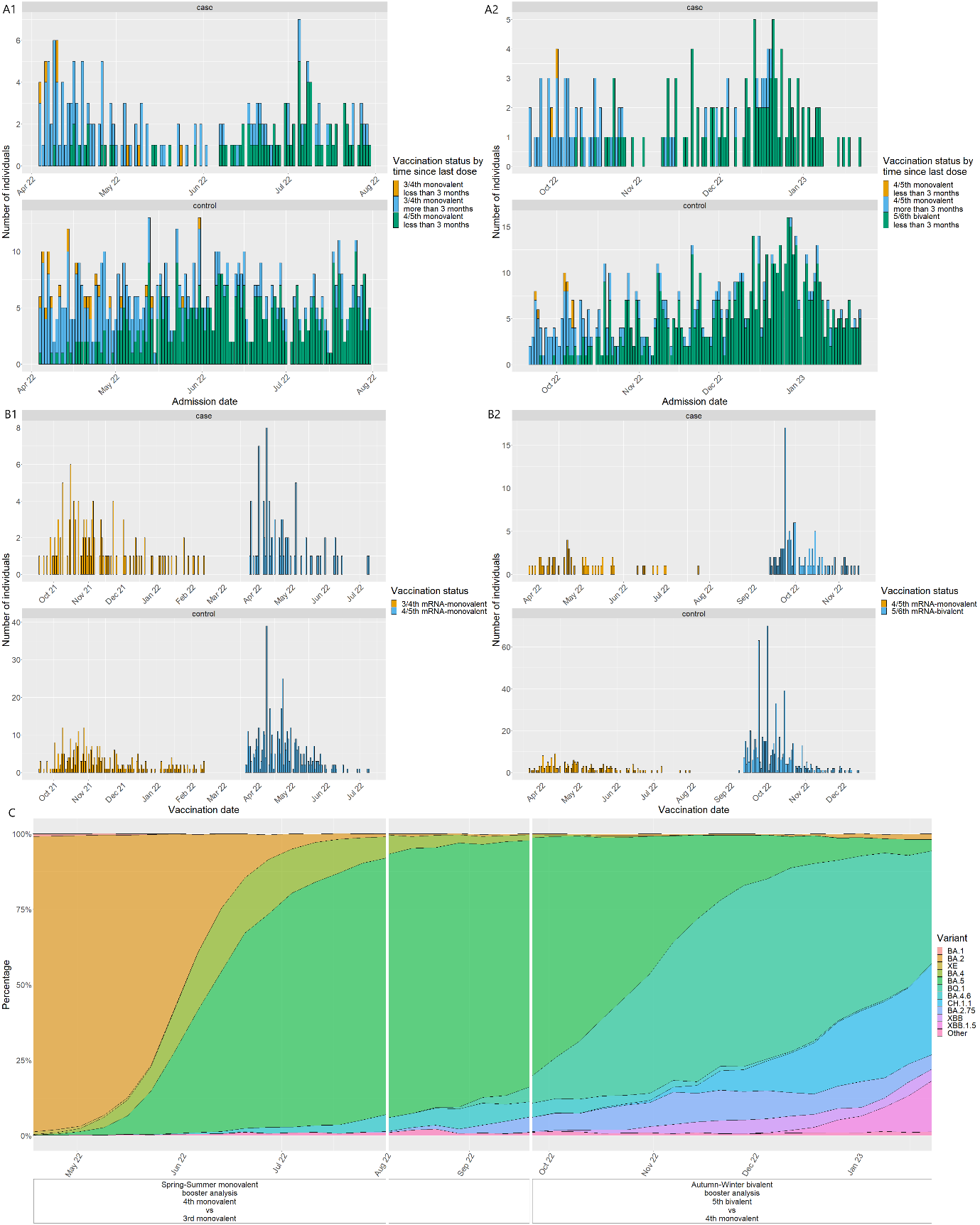

(A) The number of individuals in each cohort by vaccination status, by admission date. (B) The number of individuals in each cohort by vaccination status, by vaccination date. (C) Prevalence of different genomic variants of SARS-CoV-2 in available sequenced cases for England identified over time, as made available by UK Health Security Agency (UKHSA) (<https://assets.publishing.service.gov.uk/government/uploads/system/uploads/attachment_data/file/1152143/variant-technical-briefing-52-21-april-2023.pdf>)

**Supplementary Data 3 (S3): Admission characteristics of study participants by vaccine type [4^th^ dose – 3^rd^ dose monovalent mRNA vaccines, ≥75 years admitted between 04/04/22 and 30/07/22]**

| **Characteristic** | **4th mRNA-monovalent vaccine***  **(N = 491)** | **3rd mRNA-monovalent vaccine***  **(N = 373)** | **P-value** |
| --- | --- | --- | --- |
| Cohort |  |  | <0.001 |
| Case | 78 (16%) | 104 (28%) |  |
| Control | 413 (84%) | 269 (72%) |  |
| Vaccine brand^#^ |  |  | <0.001 |
| Moderna SpikeVax® | 285 (58%) | 39 (10%) |  |
| Pfizer Comirnaty® | 206 (42%) | 334 (90%) |  |
| Time since 3rd dose - - median days (IQR) | 224 (199, 249) | 174 (156, 196) | <0.001 |
| Time since 2nd dose - median days (IQR) | 430 (406, 457) | 381 (363, 411) |  |
| Time since last dose - - median days (IQR) | 44 (22, 68) | 174 (156, 196) | <0.001 |
| Months since last dose |  |  | <0.001 |
| ≤ 3months | 491 (100%) | 7 (1.9%) |  |
| > 3months | 0 (0%) | 366 (98%) |  |
| Age - median years (IQR) | 85 (80, 90) | 84 (79, 89) | 0.2 |
| Sex |  |  | 0.6 |
| Male | 232 (47%) | 183 (49%) |  |
| Female | 259 (53%) | 190 (51%) |  |
| LTCF Resident | 72 (15%) | 34 (9.1%) | 0.016 |
| Ethnicity |  |  | >0.9 |
| White British | 387 (79%) | 298 (80%) |  |
| Other | 22 (4.5%) | 16 (4.3%) |  |
| Unknown | 82 (17%) | 59 (16%) |  |
| IMD – median (IQR) | 5 (4, 8) | 5 (4, 8) | 0.5 |
| Unknown | 9 | 1 |  |
| Smoking |  |  | 0.061 |
| Current | 17 (3.5%) | 28 (7.5%) |  |
| Ex-smoker | 299 (61%) | 209 (56%) |  |
| Non-smoker | 151 (31%) | 117 (31%) |  |
| Unknown | 24 (4.9%) | 19 (5.1%) |  |
| Comorbidity scores |  |  |  |
| Rockwood Frailty scale |  |  | 0.7 |
| 1-4 | 92 (30%) | 81 (32%) |  |
| 5-9 | 214 (70%) | 175 (68%) |  |
| Unknown | 185 | 117 |  |
| CCI – median (IQR) | 5 (4, 6) | 5 (4, 7) | 0.3 |
| Respiratory |  |  |  |
| Any | 219 (45%) | 150 (40%) | 0.2 |
| COPD | 157 (32%) | 110 (29%) | 0.5 |
| Asthma | 51 (10%) | 34 (9.1%) | 0.6 |
| Other⁑ | 51 (10%) | 35 (9.4%) | 0.6 |
| Cardiovascular |  |  |  |
| Any | 284 (58%) | 202 (54%) | 0.3 |
| IHD | 90 (18%) | 65 (17%) | 0.8 |
| AF | 156 (32%) | 121 (32%) | 0.9 |
| CCF | 128 (26%) | 93 (25%) | 0.8 |
| Diabetes |  |  |  |
| Any | 84 (17%) | 104 (28%) | <0.001 |
| Type 1 | 0 | 0 |  |
| Type 2 | 84 (100%) | 104 (100%) |  |
| Neurological |  |  |  |
| Dementia | 67 (14%) | 50 (13%) | >0.9 |
| Cognitive impairment | 29 (5.9%) | 23 (6.2%) | 0.9 |
| CVA | 50 (10%) | 41 (11%) | 0.7 |
| TIA | 36 (7.3%) | 45 (12%) | 0.025 |
| Other neurological disease† | 20 (4.1%) | 23 (6.2%) | 0.2 |
| Immunodeficiency |  |  |  |
| CTD | 51 (10%) | 35 (9.4%) | 0.6 |
| HIV | 0 (0%) | 0 (0%) |  |
| Other immunodeficiency | 39 (7.9%) | 28 (7.5%) | 0.9 |
| Oncology |  |  |  |
| Solid organ cancer | 53 (11%) | 34 (9.1%) | 0.4 |
| Haematological malignancy | 6 (1.2%) | 4 (1.1%) | >0.9 |
| Renal disease‡ |  |  | 0.3 |
| None | 305 (62%) | 214 (57%) |  |
| Mild | 167 (34%) | 138 (37%) |  |
| Moderate/severe | 19 (3.9%) | 21 (5.6%) |  |

Data are N (%) unless otherwise stated.

*4th dose of monovalent mRNA vaccine after 21/03/22 and less than 3 months prior to admission.

3rd dose of monovalent mRNA vaccine between 16/09/21 and 14/02/22.

#Refers to vaccine brand of the last dose received (4th or 3rd dose). Prior to last dose, any vaccine combination is considered.

⁑Includes bronchiectasis, pulmonary fibrosis, and other chronic respiratory conditions.
†Includes Parkinson’s disease, Huntingdon’s disease, and other chronic neurological conditions.
‡Mild is CKD stage 1–3; moderate or severe is CKD stage 4–5, end-stage renal failure, or dialysis dependence.

AF, atrial fibrillation; CCF, congestive cardiac failure; CCI, Charlson Comorbidity Index; COPD, chronic obstructive pulmonary disease; CKD; chronic kidney disease; CVA, cerebrovascular accident; IHD, ischaemic heart disease; IMD, Index of multiple deprivation; IQR, interquartile range; LTCF, long-term care facility; TIA, transient ischaemic attack

**Supplementary Data 4 (S4): Admission characteristics of study participants by vaccine type [5^th^ dose BA.1/ancestral mRNA bivalent - 4^th^ dose monovalent vaccines, ≥75 years admitted between 21/09/22 and 23/01/23]**

| **Characteristic** | **5th mRNA-bivalent vaccine***  **(N = 672)** | **4th mRNA-monovalent vaccine ***  **(N = 212)** | **P-value** |
| --- | --- | --- | --- |
| Cohort |  |  | 0.002 |
| Case | 100 (15%) | 52 (25%) |  |
| Control | 572 (85%) | 160 (75%) |  |
| Vaccine brand^#^ |  |  | 0.003 |
| Moderna SpikeVax® | 455 (68%) | 119 (56%) |  |
| Pfizer Comirnaty® | 217 (32%) | 93 (44%) |  |
| Time since 4th dose - median days (IQR) | 223 (194, 250) | 166 (147, 190) | <0.001 |
| Time since 3rd dose - median days (IQR) | 402 (376, 425) | 350 (333, 377) | <0.001 |
| Time since 2nd dose - median days (IQR) | 609 (582, 628) | 553 (532, 597) | <0.001 |
| Time since last dose - median days (IQR) | 59 (32, 76) | 166 (147, 190) | <0.001 |
| Months since last dose |  |  | <0.001 |
| ≤ 3months | 672 (100%) | 7 (3.3%) |  |
| > 3months | 0 (0%) | 205 (97%) |  |
| Age - median years (IQR) | 84 (80, 89) | 86 (81, 89) | 0.15 |
| Sex |  |  | 0.8 |
| Male | 304 (45%) | 99 (47%) |  |
| Female | 368 (55%) | 113 (53%) |  |
| LTCF Resident | 81 (12%) | 24 (11%) | >0.9 |
| Ethnicity |  |  | 0.5 |
| White British | 529 (79%) | 163 (77%) |  |
| Other | 11 (1.6%) | 6 (2.8%) |  |
| Unknown | 132 (20%) | 43 (20%) |  |
| IMD – median (IQR) | 6 (4, 9) | 6 (4, 9) | 0.5 |
| Unknown | 9 | 2 |  |
| Smoking |  |  | 0.5 |
| Current | 409 (61%) | 129 (61%) |  |
| Ex-smoker | 184 (27%) | 65 (31%) |  |
| Non-smoker | 40 (6.0%) | 10 (4.7%) |  |
| Unknown | 39 (5.8%) | 8 (3.8%) |  |
| Comorbidity scores |  |  |  |
| Rockwood Frailty scale |  |  | 0.11 |
| 1-4 | 246 (46%) | 68 (39%) |  |
| 5-9 | 286 (54%) | 105 (61%) |  |
| Unknown | 140 | 39 |  |
| CCI – median (IQR) | 5 (4, 6) | 5 (5, 6) | 0.2 |
| Respiratory |  |  |  |
| Any | 285 (42%) | 96 (45%) | 0.5 |
| COPD | 185 (28%) | 71 (33%) | 0.10 |
| Asthma | 87 (13%) | 20 (9.4%) | 0.2 |
| Other⁑ | 61 (9.1%) | 21 (9.9%) | 0.7 |
| Cardiovascular |  |  |  |
| Any | 365 (54%) | 128 (60%) | 0.13 |
| IHD | 105 (16%) | 36 (17%) | 0.7 |
| AF | 205 (31%) | 84 (40%) | 0.015 |
| CCF | 159 (24%) | 51 (24%) | >0.9 |
| Diabetes |  |  |  |
| Any | 120 (18%) | 45 (21%) | 0.3 |
| Type 1 | 1 (0.8%) | 0 (0%) | >0.9 |
| Type 2 | 119 (99%) | 45 (100%) |  |
| Neurological |  |  |  |
| Dementia | 83 (12%) | 24 (11%) | 0.8 |
| Cognitive impairment | 50 (7.4%) | 24 (11%) | 0.087 |
| CVA | 59 (8.8%) | 26 (12%) | 0.14 |
| TIA | 62 (9.2%) | 20 (9.4%) | 0.9 |
| Other neurological disease† | 33 (4.9%) | 12 (5.7%) | 0.7 |
| Immunodeficiency |  |  |  |
| CTD | 63 (9.4%) | 16 (7.5%) | 0.5 |
| HIV | 0 (0%) | 1 (0.5%) | 0.2 |
| Other immunodeficiency | 99 (15%) | 24 (11%) | 0.3 |
| Oncology |  |  |  |
| Solid organ cancer | 60 (8.9%) | 17 (8.0%) | 0.8 |
| Haematological malignancy | 21 (3.1%) | 4 (1.9%) | 0.5 |
| Renal disease‡ |  |  | 0.5 |
| None | 353 (53%) | 121 (57%) |  |
| Mild | 287 (43%) | 84 (40%) |  |
| Moderate/severe | 32 (4.8%) | 7 (3.3%) |  |

Data are N (%) unless otherwise stated.

*5^th^ dose of BA.1/ancestral mRNA bivalent mRNA vaccine after 07/09/22 and less than 3 months prior to admission

4^th^ dose of monovalent mRNA vaccine between 21/03/22 and 07/08/22.

#Refers to vaccine brand of the last dose received (5^th^ or 4^th^ dose). Prior to last dose, any vaccine combination is considered.

⁑Includes bronchiectasis, pulmonary fibrosis, and other chronic respiratory conditions.
†Includes Parkinson’s disease, Huntingdon’s disease, and other chronic neurological conditions.
‡Mild is CKD stage 1–3; moderate or severe is CKD stage 4–5, end-stage renal failure, or dialysis dependence.

AF, atrial fibrillation; CCF, congestive cardiac failure; CCI, Charlson Comorbidity Index; COPD, chronic obstructive pulmonary disease; CKD; chronic kidney disease; CVA, cerebrovascular accident; IHD, ischaemic heart disease; IMD, Index of multiple deprivation; IQR, interquartile range; LTCF, long-term care facility; TIA, transient ischaemic attack

**Supplementary Data 5 (S5): Admission characteristics of study participants [4/5^th^ dose – 3/4^th^ dose monovalent mRNA vaccines, ≥75 years admitted between 04/04/22 and 30/07/22, including vaccinated with 3 primary doses]**

| **Characteristic** | **Cases**  **SARS-CoV-2 positive**  **(N = 199)** | **Controls**  **SARS-CoV-2 negative**  **(N = 724)** | ***P*-value** |
| --- | --- | --- | --- |
| Vaccination status* |  |  | <0.001 |
| 4/5th mRNA-monovalent | 83 (42%) | 419 (58%) |  |
| 3/4th mRNA-monovalent | 116 (58%) | 305 (42%) |  |
| Vaccine brand^#^ |  |  | 0.011 |
| Moderna SpikeVax® | 62 (31%) | 298 (41%) |  |
| Pfizer Comirnaty® | 137 (69%) | 426 (59%) |  |
| Time since 3/4th dose - median days (IQR) | 187 (160, 230) | 202 (168, 235) | 0.011 |
| Time since 2/3rd dose - median days (IQR) | 390 (361, 441) | 409 (378, 442) | <0.001 |
| Time since last dose - median days (IQR) | 135 (65, 174) | 74 (37, 163) | <0.001 |
| Months since last dose |  |  | <0.001 |
| ≤ 3months | 90 (45%) | 441 (61%) |  |
| > 3months | 109 (55%) | 283 (39%) |  |
| Primary vaccination regime |  |  | 0.2 |
| 2-dose | 182 (91%) | 682 (94%) |  |
| 3-dose | 17 (8.5%) | 42 (5.8%) |  |
| Age - median years (IQR) | 84 (79, 90) | 84 (79, 89) | 0.6 |
| Sex |  |  | 0.3 |
| Male | 106 (53%) | 351 (48%) |  |
| Female | 93 (47%) | 373 (52%) |  |
| LTCF Resident | 13 (6.5%) | 94 (13%) | 0.012 |
| Ethnicity |  |  | 0.007 |
| White British | 161 (81%) | 575 (79%) |  |
| Other | 15 (7.5%) | 24 (3.3%) |  |
| Unknown | 23 (12%) | 125 (17%) |  |
| IMD – median (IQR) | 5 (4, 8) | 6 (4, 8) | >0.9 |
| Unknown | 3 | 7 |  |
| Smoking |  |  | 0.067 |
| Current | 13 (6.5%) | 34 (4.7%) |  |
| Ex-smoker | 106 (53%) | 437 (60%) |  |
| Non-smoker | 74 (37%) | 215 (30%) |  |
| Unknown | 6 (3.0%) | 38 (5.2%) |  |
| Comorbidity scores |  |  |  |
| Rockwood Frailty scale |  |  | 0.3 |
| 1-4 | 50 (35%) | 138 (30%) |  |
| 5-9 | 91 (65%) | 315 (70%) |  |
| Unknown | 58 | 271 |  |
| CCI – median (IQR) | 5 (4, 6) | 5 (5, 6) | >0.9 |
| Respiratory |  |  |  |
| Any | 74 (37%) | 335 (46%) | 0.024 |
| COPD | 47 (24%) | 246 (34%) | 0.006 |
| Asthma | 19 (9.5%) | 76 (10%) | 0.8 |
| Other⁑ | 22 (11%) | 84 (12%) | >0.9 |
| Cardiovascular |  |  |  |
| Any | 108 (54%) | 413 (57%) | 0.5 |
| IHD | 32 (16%) | 130 (18%) | 0.6 |
| AF | 63 (32%) | 233 (32%) | >0.9 |
| CCF | 47 (24%) | 187 (26%) | 0.6 |
| Diabetes |  |  |  |
| Any | 47 (24%) | 155 (21%) | 0.5 |
| Type 1 | 0 (0%) | 1 (0.6%) | >0.9 |
| Type 2 | 47 (100%) | 154 (99%) |  |
| Neurological |  |  |  |
| Dementia | 24 (12%) | 95 (13%) | 0.8 |
| Cognitive impairment | 15 (7.5%) | 38 (5.2%) | 0.2 |
| CVA | 18 (9.0%) | 76 (10%) | 0.6 |
| TIA | 15 (7.5%) | 73 (10%) | 0.3 |
| Other neurological disease† | 12 (6.0%) | 34 (4.7%) | 0.5 |
| Immunodeficiency |  |  |  |
| CTD | 24 (12%) | 75 (10%) | 0.5 |
| HIV | 1 (0.5%) | 0 (0%) | 0.2 |
| Other immunodeficiency | 19 (9.5%) | 60 (8.3%) | 0.6 |
| Oncology |  |  |  |
| Solid organ cancer | 20 (10%) | 75 (10%) | >0.9 |
| Haematological malignancy | 7 (3.5%) | 14 (1.9%) | 0.2 |
| Renal disease‡ |  |  | 0.5 |
| None | 117 (59%) | 433 (60%) |  |
| Mild | 69 (35%) | 258 (36%) |  |
| Moderate/severe | 13 (6.5%) | 33 (4.6%) |  |

Data are N (%) unless otherwise stated.

*Vaccinated individuals who have received a 4/5^th^ dose of monovalent mRNA vaccine after 21/03/22 and less than 3 months prior to admission or a 3/4^th^ dose of monovalent mRNA vaccine between 16/09/21 and 14/02/22.

#Refers to vaccine brand of the last dose received (4/5^th^ or 3/4^th^ dose). Prior to last dose, any vaccine combination is considered.

⁑Includes bronchiectasis, pulmonary fibrosis, and other chronic respiratory conditions.
†Includes Parkinson’s disease, Huntingdon’s disease, and other chronic neurological conditions.
‡Mild is CKD stage 1–3; moderate or severe is CKD stage 4–5, end-stage renal failure, or dialysis dependence.

AF, atrial fibrillation; CCF, congestive cardiac failure; CCI, Charlson Comorbidity Index; COPD, chronic obstructive pulmonary disease; CKD; chronic kidney disease; CVA, cerebrovascular accident; IHD, ischaemic heart disease; IMD, Index of multiple deprivation; IQR, interquartile range; LTCF, long-term care facility; TIA, transient ischaemic attack

**Supplementary Data 6 (S6): Admission characteristics of study participants by vaccine type [4/5^th^ dose – 3/4^th^ dose monovalent mRNA vaccines, ≥75 years admitted between 04/04/22 and 30/07/22, including vaccinated with 3 primary doses]**

| **Characteristic** | **4/5th mRNA-monovalent vaccine***  **(N = 502)** | **3/4th mRNA-monovalent vaccine***  **(N = 421)** | **P-value** |
| --- | --- | --- | --- |
| Cohort |  |  | <0.001 |
| Case | 83 (17%) | 116 (28%) |  |
| Control | 419 (83%) | 305 (72%) |  |
| Vaccine brand^#^ |  |  | <0.001 |
| Moderna SpikeVax® | 292 (58%) | 68 (16%) |  |
| Pfizer Comirnaty® | 210 (42%) | 353 (84%) |  |
| Time since 3/4th dose - median days (IQR) | 224 (197, 247) | 170 (148, 192) | <0.001 |
| Time since 2/3th dose - median days (IQR) | 430 (404, 457) | 376 (353, 405) | <0.001 |
| Time since last dose - median days (IQR) | 44 (21, 68) | 170 (148, 192) | <0.001 |
| Months since last dose |  |  | <0.001 |
| ≤ 3months | 502 (100%) | 29 (6.9%) |  |
| > 3months | 0 (0%) | 392 (93%) |  |
| Primary vaccination regime |  |  | <0.001 |
| 2-dose | 491 (98%) | 373 (89%) |  |
| 3-dose | 11 (2.2%) | 48 (11%) |  |
| Age - median years (IQR) | 85 (80, 90) | 83 (79, 88) | 0.044 |
| Sex |  |  | 0.3 |
| Male | 240 (48%) | 217 (52%) |  |
| Female | 262 (52%) | 204 (48%) |  |
| LTCF Resident | 72 (14%) | 35 (8.3%) | 0.005 |
| Ethnicity |  |  | 0.8 |
| White British | 396 (79%) | 340 (81%) |  |
| Other | 22 (4.4%) | 17 (4.0%) |  |
| Unknown | 84 (17%) | 64 (15%) |  |
| IMD – median (IQR) | 5 (4, 8) | 6 (4, 9) | 0.2 |
| Unknown | 9 | 1 |  |
| Smoking |  |  | 0.059 |
| Current | 17 (3.4%) | 30 (7.1%) |  |
| Ex-smoker | 307 (61%) | 236 (56%) |  |
| Non-smoker | 154 (31%) | 135 (32%) |  |
| Unknown | 24 (4.8%) | 20 (4.8%) |  |
| Comorbidity scores |  |  |  |
| Rockwood Frailty scale |  |  | 0.5 |
| 1-4 | 95 (30%) | 93 (33%) |  |
| 5-9 | 218 (70%) | 188 (67%) |  |
| Unknown | 189 | 140 |  |
| CCI – median (IQR) | 5 (5, 6) | 5 (4, 7) | 0.2 |
| Respiratory |  |  |  |
| Any | 227 (45%) | 182 (43%) | 0.6 |
| COPD | 160 (32%) | 133 (32%) | >0.9 |
| Asthma | 54 (11%) | 41 (9.7%) | 0.7 |
| Other⁑ | 56 (11%) | 50 (12%) | 0.8 |
| Cardiovascular |  |  |  |
| Any | 292 (58%) | 229 (54%) | 0.3 |
| IHD | 90 (18%) | 72 (17%) | 0.8 |
| AF | 159 (32%) | 137 (33%) | 0.8 |
| CCF | 134 (27%) | 100 (24%) | 0.3 |
| Diabetes |  |  |  |
| Any | 86 (17%) | 116 (28%) | <0.001 |
| Type 1 | 0 (0%) | 1 (0.9%) | >0.9 |
| Type 2 | 86 (100%) | 115 (99%) |  |
| Neurological |  |  |  |
| Dementia | 67 (13%) | 52 (12%) | 0.7 |
| Cognitive impairment | 29 (5.8%) | 24 (5.7%) | >0.9 |
| CVA | 51 (10%) | 43 (10%) | >0.9 |
| TIA | 38 (7.6%) | 50 (12%) | 0.032 |
| Other neurological disease† | 20 (4.0%) | 26 (6.2%) | 0.13 |
| Immunodeficiency |  |  |  |
| CTD | 52 (10%) | 47 (11%) | 0.7 |
| HIV | 0 (0%) | 1 (0.2%) | 0.5 |
| Other immunodeficiency | 40 (8.0%) | 39 (9.3%) | 0.6 |
| Oncology |  |  |  |
| Solid organ cancer | 54 (11%) | 41 (9.7%) | 0.7 |
| Haematological malignancy | 8 (1.6%) | 13 (3.1%) | 0.2 |
| Renal disease‡ |  |  | 0.13 |
| None | 312 (62%) | 238 (57%) |  |
| Mild | 170 (34%) | 157 (37%) |  |
| Moderate/severe | 20 (4.0%) | 26 (6.2%) |  |

Data are N (%) unless otherwise stated.

*4/5th dose of monovalent mRNA vaccine after 21/03/22 and less than 3 months prior to admission.

3/4th dose of monovalent mRNA vaccine between 16/09/21 and 14/02/22.

#Refers to vaccine brand of the last dose received (4/5^th^ or 3/4^th^ dose). Prior to last dose, any vaccine combination is considered.

⁑Includes bronchiectasis, pulmonary fibrosis, and other chronic respiratory conditions.
†Includes Parkinson’s disease, Huntingdon’s disease, and other chronic neurological conditions.
‡Mild is CKD stage 1–3; moderate or severe is CKD stage 4–5, end-stage renal failure, or dialysis dependence.

AF, atrial fibrillation; CCF, congestive cardiac failure; CCI, Charlson Comorbidity Index; COPD, chronic obstructive pulmonary disease; CKD; chronic kidney disease; CVA, cerebrovascular accident; IHD, ischaemic heart disease; IMD, Index of multiple deprivation; IQR, interquartile range; LTCF, long-term care facility; TIA, transient ischaemic attack

**Supplementary Data 7 (S7): Admission characteristics of study participants [5/6^th^ dose BA.1/ancestral mRNA bivalent – 4/5^th^ dose monovalent vaccines, ≥75 years admitted between 21/09/22 and 23/01/23, including vaccinated with 3 primary doses]**

| **Characteristic** | **Cases**  **SARS-CoV-2 positive**  **(N = 158)** | **Controls**  **SARS-CoV-2 negative**  **(N = 786)** | ***P*-value** |
| --- | --- | --- | --- |
| Vaccination status* |  |  | 0.002 |
| 5th mRNA-bivalent | 103 (65%) | 610 (78%) |  |
| 4th mRNA-monovalent | 55 (35%) | 176 (22%) |  |
| Vaccine brand^#^ |  |  | 0.2 |
| Moderna SpikeVax® | 95 (60%) | 517 (66%) |  |
| Pfizer Comirnaty® | 63 (40%) | 269 (34%) |  |
| Time since 4/5th dose - median days (IQR) | 204 (164, 237) | 207 (176, 242) | 0.3 |
| Time since 3/4th dose - median days (IQR) | 384 (346, 418) | 390 (355, 419) | 0.6 |
| Time since 2/3rd dose - median days (IQR) | 592 (548, 620) | 597 (560, 623) | 0.5 |
| Time since last dose - median days (IQR) | 75 (52, 144) | 67 (40, 97) | 0.03 |
| Months since last dose |  |  | 0.001 |
| ≤ 3months | 105 (66%) | 618 (79%) |  |
| > 3months | 53 (34%) | 168 (21%) |  |
| Primary vaccination regime |  |  | 0.2 |
| 2-dose | 152 (96%) | 732 (93%) |  |
| 3-dose | 6 (3.8%) | 54 (6.9%) |  |
| Age - median years (IQR) | 84 (80, 89) | 85 (80, 89) | 0.4 |
| Sex |  |  | 0.2 |
| Male | 80 (51%) | 351 (45%) |  |
| Female | 78 (49%) | 435 (55%) |  |
| LTCF Resident | 19 (12%) | 89 (11%) | 0.8 |
| Ethnicity |  |  | 0.061 |
| White British | 113 (72%) | 627 (80%) |  |
| Other | 4 (2.5%) | 13 (1.7%) |  |
| Unknown | 41 (26%) | 146 (19%) |  |
| IMD – median (IQR) | 6 (4, 9) | 6 (4, 9) | 0.6 |
| Unknown | 2 | 10 |  |
| Smoking |  |  | 0.003 |
| Current | 2 (1.3%) | 53 (6.7%) |  |
| Ex-smoker | 89 (56%) | 482 (61%) |  |
| Non-smoker | 55 (35%) | 215 (27%) |  |
| Unknown | 12 (7.6%) | 36 (4.6%) |  |
| Comorbidity scores |  |  |  |
| Rockwood Frailty scale |  |  | 0.14 |
| 1-4 | 47 (39%) | 290 (46%) |  |
| 5-9 | 75 (61%) | 336 (54%) |  |
| Unknown | 36 | 160 |  |
| CCI – median (IQR) | 5 (4, 6) | 5 (4, 7) | 0.7 |
| Respiratory |  |  |  |
| Any | 61 (39%) | 346 (44%) | 0.2 |
| COPD | 39 (25%) | 237 (30%) | 0.2 |
| Asthma | 18 (11%) | 94 (12%) | >0.9 |
| Other⁑ | 10 (6.3%) | 79 (10%) | 0.2 |
| Cardiovascular |  |  |  |
| Any | 87 (55%) | 440 (56%) | 0.9 |
| IHD | 31 (20%) | 123 (16%) | 0.2 |
| AF | 50 (32%) | 258 (33%) | 0.9 |
| CCF | 31 (20%) | 190 (24%) | 0.3 |
| Diabetes |  |  |  |
| Any | 26 (16%) | 157 (20%) | 0.4 |
| Type 1 | 0 (0%) | 1 (0.6%) | >0.9 |
| Type 2 | 26 (100%) | 156 (99%) |  |
| Neurological |  |  |  |
| Dementia | 27 (17%) | 83 (11%) | 0.029 |
| Cognitive impairment | 18 (11%) | 58 (7.4%) | 0.11 |
| CVA | 21 (13%) | 69 (8.8%) | 0.10 |
| TIA | 18 (11%) | 66 (8.4%) | 0.2 |
| Other neurological disease† | 11 (7.0%) | 36 (4.6%) | 0.2 |
| Immunodeficiency |  |  |  |
| CTD | 18 (11%) | 76 (9.7%) | 0.6 |
| HIV | 0 (0%) | 1 (0.1%) | >0.9 |
| Other immunodeficiency | 23 (15%) | 120 (15%) | >0.9 |
| Oncology |  |  |  |
| Solid organ cancer | 12 (7.6%) | 77 (9.8%) | 0.5 |
| Haematological malignancy | 4 (2.5%) | 32 (4.1%) | 0.5 |
| Renal disease‡ |  |  | 0.5 |
| None | 91 (58%) | 417 (53%) |  |
| Mild | 62 (39%) | 330 (42%) |  |
| Moderate/severe | 5 (3.2%) | 39 (5.0%) |  |

Data are N (%) unless otherwise stated.

*Vaccinated individuals who have received a 5/6^th^ dose of BA.1/ancestral mRNA bivalent mRNA vaccine after 07/09/22 and less than 3 months prior to admission or a 4/5^th^ dose of monovalent mRNA vaccine between 21/03/22 and 07/08/22.

#Refers to vaccine brand of the last dose received (5/6^th^ or 4/5^th^ dose). Prior to last dose, any vaccine combination is considered.

⁑Includes bronchiectasis, pulmonary fibrosis, and other chronic respiratory conditions.
†Includes Parkinson’s disease, Huntingdon’s disease, and other chronic neurological conditions.
‡Mild is CKD stage 1–3; moderate or severe is CKD stage 4–5, end-stage renal failure, or dialysis dependence.

AF, atrial fibrillation; CCF, congestive cardiac failure; CCI, Charlson Comorbidity Index; COPD, chronic obstructive pulmonary disease; CKD; chronic kidney disease; CVA, cerebrovascular accident; IHD, ischaemic heart disease; IMD, Index of multiple deprivation; IQR, interquartile range; LTCF, long-term care facility; TIA, transient ischaemic attack

**Supplementary Data 8 (S8): Admission characteristics of study participants by vaccine type [5/6^th^ dose BA.1/ancestral mRNA bivalent – 4/5^th^ dose monovalent vaccines, ≥75 years admitted between 21/09/22 and 23/01/23, including vaccinated with 3 primary doses]**

| **Characteristic** | **5/6^th^ mRNA-bivalent vaccine***  **(N = 713)** | **4/5^th^ mRNA-monovalent vaccine ***  **(N = 231)** | **P-value** |
| --- | --- | --- | --- |
| Cohort |  |  | 0.002 |
| Case | 103 (14%) | 55 (24%) |  |
| Control | 610 (86%) | 176 (76%) |  |
| Vaccine brand^#^ |  |  | 0.022 |
| Moderna SpikeVax® | 477 (67%) | 135 (58%) |  |
| Pfizer Comirnaty® | 236 (33%) | 96 (42%) |  |
| Time since 4/5^th^ dose - median days (IQR) | 219 (191, 249) | 164 (144, 188) | <0.001 |
| Time since 3/4th dose - median days (IQR) | 400 (371, 424) | 346 (327, 373) | <0.001 |
| Time since 2/3rd dose - median days (IQR) | 606 (577, 627) | 549 (527, 590) | <0.001 |
| Time since last dose - median days (IQR) | 58 (32, 76) | 164 (144, 188) | <0.001 |
| Months since last dose |  |  | <0.001 |
| ≤ 3months | 713 (100%) | 10 (4.3%) |  |
| > 3months | 0 (0%) | 221 (96%) |  |
| Primary vaccination regime |  |  | 0.2 |
| 2-dose | 672 (94%) | 212 (92%) |  |
| 3-dose | 41 (5.8%) | 19 (8.2%) |  |
| Age – median years (IQR) | 84 (80, 89) | 85 (80, 89) | 0.3 |
| Sex |  |  | 0.8 |
| Male | 324 (45%) | 107 (46%) |  |
| Female | 389 (55%) | 124 (54%) |  |
| LTCF Resident | 83 (12%) | 25 (11%) | 0.8 |
| Ethnicity |  |  | 0.6 |
| White British | 560 (79%) | 180 (78%) |  |
| Other | 11 (1.5%) | 6 (2.6%) |  |
| Unknown | 142 (20%) | 45 (19%) |  |
| IMD – median (IQR) | 6 (4, 9) | 6 (4, 9) | 0.5 |
| Unknown | 10 | 2 |  |
| Smoking |  |  | 0.3 |
| Current | 44 (6.2%) | 11 (4.8%) |  |
| Ex-smoker | 434 (61%) | 137 (59%) |  |
| Non-smoker | 195 (27%) | 75 (32%) |  |
| Unknown | 40 (5.6%) | 8 (3.5%) |  |
| Comorbidity scores |  |  |  |
| Rockwood Frailty scale |  |  | 0.13 |
| 1-4 | 263 (47%) | 74 (40%) |  |
| 5-9 | 300 (53%) | 111 (60%) |  |
| Unknown | 150 | 46 |  |
| CCI – median (IQR) | 5 (4, 6) | 5 (5, 6) | 0.3 |
| Respiratory |  |  |  |
| Any | 303 (42%) | 104 (45%) | 0.5 |
| COPD | 200 (28%) | 76 (33%) | 0.2 |
| Asthma | 91 (13%) | 21 (9.1%) | 0.2 |
| Other⁑ | 64 (9.0%) | 25 (11%) | 0.4 |
| Cardiovascular |  |  |  |
| Any | 388 (54%) | 139 (60%) | 0.13 |
| IHD | 114 (16%) | 40 (17%) | 0.7 |
| AF | 220 (31%) | 88 (38%) | 0.044 |
| CCF | 168 (24%) | 53 (23%) | >0.9 |
| Diabetes |  |  |  |
| Any | 134 (19%) | 49 (21%) | 0.4 |
| Type 1 | 1 (0.7%) | 0 (0%) | >0.9 |
| Type 2 | 133 (99%) | 49 (100%) |  |
| Neurological |  |  |  |
| Dementia | 86 (12%) | 24 (10%) | 0.6 |
| Cognitive impairment | 51 (7.2%) | 25 (11%) | 0.094 |
| CVA | 64 (9.0%) | 26 (11%) | 0.3 |
| TIA | 64 (9.0%) | 20 (8.7%) | >0.9 |
| Other neurological disease† | 35 (4.9%) | 12 (5.2%) | 0.9 |
| Immunodeficiency |  |  |  |
| CTD | 71 (10.0%) | 23 (10.0%) | >0.9 |
| HIV | 0 (0%) | 1 (0.4%) | 0.2 |
| Other immunodeficiency | 110 (15%) | 33 (14%) | 0.8 |
| Oncology |  |  |  |
| Solid organ cancer | 68 (9.5%) | 21 (9.1%) | 0.9 |
| Haematological malignancy | 28 (3.9%) | 8 (3.5%) | 0.8 |
| Renal disease‡ |  |  | 0.3 |
| None | 374 (52%) | 134 (58%) |  |
| Mild | 303 (42%) | 89 (39%) |  |
| Moderate/severe | 36 (5.0%) | 8 (3.5%) |  |

Data are N (%) unless otherwise stated.

*5/6^th^ dose of BA.1/ancestral mRNA bivalent mRNA vaccine after 07/09/22 and less than 3 months prior to admission

4/5^th^ dose of monovalent mRNA vaccine between 21/03/22 and 07/08/22.

#Refers to vaccine brand of the last dose received (5/6^th^ or 4/5^th^ dose). Prior to last dose, any vaccine combination is considered.

⁑Includes bronchiectasis, pulmonary fibrosis, and other chronic respiratory conditions.
†Includes Parkinson’s disease, Huntingdon’s disease, and other chronic neurological conditions.
‡Mild is CKD stage 1–3; moderate or severe is CKD stage 4–5, end-stage renal failure, or dialysis dependence.

AF, atrial fibrillation; CCF, congestive cardiac failure; CCI, Charlson Comorbidity Index; COPD, chronic obstructive pulmonary disease; CKD; chronic kidney disease; CVA, cerebrovascular accident; IHD, ischaemic heart disease; IMD, Index of multiple deprivation; IQR, interquartile range; LTCF, long-term care facility; TIA, transient ischaemic attack

**Supplementary Data 9 (S9): Relative vaccine effectiveness of 4/5^th^ dose mRNA monovalent vaccines against hospitalisation, compared to 3/4^th^ dose monovalent mRNA vaccines [04/04/22-30/07/22, including vaccinated with 3 primary doses]**

| **Characteristic** | **rVE (95% CI)** | **rOR (95% CI)** | ***P*-value** |
| --- | --- | --- | --- |
| **Univariable Logistic Regression Model** | | | |
| 4/5^th^ dose of monovalent mRNA vaccine | 47.9 (28.5, 62.2) | 0.521 (0.378, 0.715) | <0.001 |
| **Multivariable Logistic Regression Model** | | | |
| 4/5^th^ dose of monovalent mRNA vaccine | 37.4 ( 2.1, 60.2 ) | 0.626 ( 0.398, 0.979 ) | 0.041 |
| Time between 3/4th dose and admission |  | 1.004 ( 0.999, 1.008 ) | 0.14 |
| Vaccine brand^*^ |  | 0.878 ( 0.587, 1.310 ) | 0.5 |
| Primary vaccination series⁑ |  | 0.508 ( 0.248, 1.063 ) | 0.066 |
| Age |  | 1.008 ( 0.980, 1.037 ) | 0.6 |
| Sex (Male) |  | 1.057 ( 0.756, 1.479 ) | 0.7 |
| CCI |  | 0.974 ( 0.879, 1.074 ) | 0.6 |
| IMD |  | 0.969 ( 0.912, 1.030 ) | 0.3 |
| LTCF Resident |  | 0.434 ( 0.217, 0.799 ) | 0.011 |
| Respiratory disease |  | 0.643 ( 0.451, 0.913 ) | 0.014 |
| Prevalence**#** |  | 1.001 ( 1.001, 1.002 ) | <0.001 |
| **Matched Conditional Logistic Regression Model†** | | | |
| 4/5th dose of monovalent mRNA vaccine | 37.9 ( 1.0, 61.0 ) | 0.621 ( 0.390, 0.990 ) | 0.045 |
| Time between 3/4th dose and admission |  | 1.004 ( 0.999, 1.008 ) | 0.12 |
| Vaccine brand |  | 0.820 ( 0.548, 1.227 ) | 0.3 |
| Primary vaccination series⁑ |  | 0.583 ( 0.284, 1.199 ) | 0.14 |
| Prevalence**#** |  | 1.001 ( 1.001, 1.002 ) | <0.001 |

CI, Confidence Interval; CCI, Charlson comorbidity index; IMD, index of multiple deprivation; rOR, relative Odds Ratio; rVE, relative vaccine effectiveness

* Vaccine brand of 4/5^th^ dose of monovalent mRNA vaccine or 3/4^th^ dose of monovalent mRNA vaccine (prior to last dose, any vaccine combination is considered), where 1= Moderna SpikeVax® and 0 = Pfizer Comirnaty®

⁑ Primary vaccination series, where 1= 2 doses before autumn-winter 2021 and 0= 3 doses before autumn-winter 2021

**#**Prevalence was calculated on a daily basis

**†**1:3 Nearest neighbour propensity score matching with replacement (propensity score estimated using logistic regression on age, sex, CCI score, IMD, LTCF residency and respiratory disease), 196 test-positive cases were matched to 588 test-negative controls with no match found for 129 controls.

**Supplementary Data 10 (S10): Relative vaccine effectiveness of 5/6^th^ dose BA.1/ancestral mRNA bivalent vaccines against hospitalisation, compared to 4/5^th^ dose monovalent mRNA vaccines [21/09/22 – 23/01/23, including vaccinated with 3 primary doses]**

| **Characteristic** | **rVE (95% CI)** | **rOR (95% CI)** | ***P*-value** |
| --- | --- | --- | --- |
| **Univariable Logistic Regression Model** | | | |
| 5/6th dose of monovalent mRNA vaccine | 46.0 (21.6, 62.5) | 0.540 (0.375, 0.784) | 0.001 |
| **Multivariable Logistic Regression Model** | | | |
| 5/6th dose of monovalent mRNA vaccine | 46.8 ( 18.9, 64.9 ) | 0.532 ( 0.351, 0.811 ) | 0.003 |
| Time between 4/5th dose and admission |  | 1.000 ( 0.996, 1.002 ) | 0.8 |
| Vaccine brand^*^ |  | 0.828 ( 0.579, 1.192 ) | 0.3 |
| Primary vaccination series⁑ |  | 1.963 ( 0.864, 5.307 ) | 0.14 |
| Age |  | 1.004 ( 0.973, 1.036 ) | 0.8 |
| Sex (Male) |  | 1.302 ( 0.913, 1.858 ) | 0.14 |
| CCI |  | 0.981 ( 0.875, 1.093 ) | 0.7 |
| IMD |  | 1.013 ( 0.952, 1.078 ) | 0.7 |
| LTCF Resident |  | 1.004 ( 0.561 1.715 ) | >0.9 |
| Respiratory disease |  | 0.794 ( 0.546, 1.146 ) | 0.2 |
| Prevalence |  | 1.006 ( 0.999, 1.014 ) | 0.086 |
| **Matched Conditional Logistic Regression Model†** | | | |
| 5/6th dose of monovalent mRNA vaccine | 44.9 ( 15.8, 64.0 ) | 0.551 ( 0.360, 0.842 ) | 0.006 |
| Time between 4/5th dose and admission |  | 1.000 ( 0.997, 1.003 ) | >0.9 |
| Vaccine brand* |  | 0.852 ( 0.591, 1.229 ) | 0.4 |
| Primary vaccination series⁑ |  | 1.970 ( 0.780, 4.976 ) | 0.2 |
| Prevalence# |  | 1.007 ( 0.999, 1.014 ) | 0.089 |

CI, Confidence Interval; CCI, Charlson comorbidity index; IMD, index of multiple deprivation; rOR, relative Odds Ratio; rVE, relative vaccine effectiveness

* Vaccine brand of 5/6^th^ dose of BA.1/ancestral mRNA bivalent mRNA vaccine or 4/5^th^ dose of monovalent mRNA vaccine (prior to last dose, any vaccine combination is considered), where 1= Moderna SpikeVax® and 0 = Pfizer Comirnaty®

⁑ Primary vaccination series, where 1= 2 doses before autumn-winter 2021 and 0= 3 doses before autumn-winter 2021

**#**Prevalence was calculated on a daily basis

**†**1:4 Nearest neighbour propensity score matching with replacement (propensity score estimated using logistic regression on age, sex, CCI score, IMD, LTCF residency and respiratory disease), 156 test-positive cases were matched to 624 test-negative controls with no match found for 152 controls.
