## Supplementary material for "Relative vaccine effectiveness (rVE) of mRNA COVID-19 boosters in people aged at least 75 years in the UK vaccination programme, during the Spring-Summer (monovalent vaccine) and Autumn-Winter 2022 (bivalent vaccine) booster campaigns: a prospective test negative case-control study": Tables

**Table 1: Admission characteristics of study participants [4^th^ dose – 3^rd^ dose monovalent mRNA vaccines, ≥75 years admitted between 04/04/22 and 30/07/22]**

| **Characteristic** | **Cases**  **SARS-CoV-2 positive**  **(N = 182)** | **Controls**  **SARS-CoV-2 negative**  **(N = 682)** | ***P*-value** |
| --- | --- | --- | --- |
| Vaccination status* |  |  | <0.001 |
| 4th mRNA-monovalent | 78 (43%) | 413 (61%) |  |
| 3rd mRNA-monovalent | 104 (57%) | 269 (39%) |  |
| Vaccine brand^#^ |  |  | 0.010 |
| Moderna SpikeVax® | 53 (29%) | 271 (40%) |  |
| Pfizer Comirnaty® | 129 (71%) | 411 (60%) |  |
| Time since 3rd dose - median days (IQR) | 191 (165, 235) | 205 (174, 237) | 0.028 |
| Time since 2nd dose - median days (IQR) | 402 (368, 444) | 412 (382, 443) | 0.002 |
| Time since last dose - median days (IQR) | 140 (65, 175) | 72 (35, 165) | <0.001 |
| Months since last dose |  |  | <0.001 |
| ≤ 3months | 79 (43%) | 419 (61%) |  |
| > 3months | 103 (57%) | 263 (39%) |  |
| Age - median years (IQR) | 85 (79, 90) | 84 (80, 89) | 0.4 |
| Sex |  |  | 0.2 |
| Male | 96 (53%) | 319 (47%) |  |
| Female | 86 (47%) | 363 (53%) |  |
| LTCF Resident | 13 (7.1%) | 93 (14%) | 0.016 |
| Ethnicity |  |  | 0.016 |
| White British | 147 (81%) | 538 (79%) |  |
| Other | 14 (7.7%) | 24 (3.5%) |  |
| Unknown | 21 (12%) | 120 (18%) |  |
| IMD – median (IQR) | 5 (4, 8) | 5 (4, 8) | >0.9 |
| Unknown | 3 | 7 |  |
| Smoking |  |  | 0.045 |
| Current | 12 (6.6%) | 33 (4.8%) |  |
| Ex-smoker | 94 (52%) | 414 (61%) |  |
| Non-smoker | 70 (38%) | 198 (29%) |  |
| Unknown | 6 (3.3%) | 37 (5.4%) |  |
| Comorbidity scores |  |  |  |
| Rockwood Frailty scale |  |  | 0.4 |
| 1-4 | 44 (34%) | 129 (30%) |  |
| 5-9 | 85 (66%) | 304 (70%) |  |
| Unknown | 53 | 249 |  |
| CCI – median (IQR) | 5 (4, 6) | 5 (4, 6) | 0.9 |
| Respiratory |  |  |  |
| Any^ | 63 (35%) | 306 (45%) | 0.014 |
| COPD | 39 (21%) | 228 (33%) | 0.002 |
| Asthma | 16 (8.8%) | 69 (10%) | 0.7 |
| Other⁑ | 16 (8.8%) | 70 (10%) | 0.7 |
| Cardiovascular |  |  |  |
| Any | 99 (54%) | 387 (57%) | 0.6 |
| IHD | 30 (16%) | 125 (18%) | 0.7 |
| AF | 59 (32%) | 218 (32%) | >0.9 |
| CCF | 42 (23%) | 179 (26%) | 0.4 |
| Diabetes |  |  |  |
| Any | 46 (25%) | 142 (21%) | 0.2 |
| Type 1 | 0 | 0 |  |
| Type 2 | 46 (100%) | 142 (100%) |  |
| Neurological |  |  |  |
| Dementia | 24 (13%) | 93 (14%) | >0.9 |
| Cognitive impairment | 15 (8.2%) | 37 (5.4%) | 0.2 |
| CVA | 18 (9.9%) | 73 (11%) | 0.9 |
| TIA | 14 (7.7%) | 67 (9.8%) | 0.5 |
| Other neurological disease† | 11 (6.0%) | 32 (4.7%) | 0.4 |
| Immunodeficiency |  |  |  |
| CTD | 19 (10%) | 67 (9.8%) | 0.8 |
| HIV | 0 (0%) | 0 (0%) |  |
| Other immunodeficiency | 15 (8.2%) | 52 (7.6%) | 0.8 |
| Oncology |  |  |  |
| Solid organ cancer | 17 (9.3%) | 70 (10%) | 0.8 |
| Haematological malignancy | 3 (1.6%) | 7 (1.0%) | 0.4 |
| Renal disease‡ |  |  | 0.2 |
| None | 105 (58%) | 414 (61%) |  |
| Mild | 64 (35%) | 241 (35%) |  |
| Moderate/severe | 13 (7.1%) | 27 (4.0%) |  |

Data are N (%) unless otherwise stated.

*Vaccinated individuals who have received a 4^th^ dose of monovalent mRNA vaccine after 21/03/22 and less than 3 months prior to admission or a 3^rd^ dose of monovalent mRNA vaccine between 16/09/21 and 14/02/22.

#Refers to vaccine brand of the last dose received (4^th^ or 3^rd^). Prior to last dose, any vaccine combination is considered.

^Includes any chronic respiratory condition on admission to hospital, such as COPD, asthma, bronchiectasis, pulmonary fibrosis or a rare lung disease
⁑Includes bronchiectasis, pulmonary fibrosis, and other rare chronic respiratory conditions.
†Includes Parkinson’s disease, Huntingdon’s disease, and other chronic neurological conditions.
‡Mild is CKD stage 1–3; moderate or severe is CKD stage 4–5, end-stage renal failure, or dialysis dependence.

AF, atrial fibrillation; CCF, congestive cardiac failure; CCI, Charlson Comorbidity Index; COPD, chronic obstructive pulmonary disease; CKD; chronic kidney disease; CVA, cerebrovascular accident; IHD, ischaemic heart disease; IMD, Index of multiple deprivation; IQR, interquartile range; LTCF, long-term care facility; TIA, transient ischaemic attack

**Table 2: Admission characteristics of study participants [5^th^ dose BA.1/ancestral mRNA bivalent - 4^th^ dose monovalent vaccines, ≥75 years admitted between 21/09/22 and 23/01/23]**

| **Characteristic** | **Cases**  **SARS-CoV-2 positive(N = 152)** | **Controls**  **SARS-CoV-2 negative**  **(N = 732)** | ***P*-value** |
| --- | --- | --- | --- |
| Vaccination status* |  |  | 0.002 |
| 5th mRNA-bivalent | 100 (66%) | 572 (78%) |  |
| 4th mRNA-monovalent | 52 (34%) | 160 (22%) |  |
| Vaccine brand^#^ |  |  | 0.3 |
| Moderna SpikeVax® | 93 (61%) | 481 (66%) |  |
| Pfizer Comirnaty® | 59 (39%) | 251 (34%) |  |
| Time since 4th dose - median days (IQR) | 205 (169, 238) | 210 (179, 244) | 0.4 |
| Time since 3rd dose - median days (IQR) | 386 (353, 418) | 394 (362, 421) | 0.3 |
| Time since 2nd dose - median days (IQR) | 595 (552, 620) | 601 (569, 625) | 0.1 |
| Time since last dose - median days (IQR) | 75 (52, 145) | 67 (40, 97) | 0.023 |
| Months since last dose |  |  | 0.001 |
| ≤ 3months | 101 (66%) | 578 (79%) |  |
| > 3months | 51 (34%) | 154 (21%) |  |
| Age - median years (IQR) | 85 (81, 89) | 85 (80, 89) | 0.6 |
| Sex |  |  | 0.13 |
| Male | 78 (51%) | 325 (44%) |  |
| Female | 74 (49%) | 407 (56%) |  |
| LTCF Resident | 18 (12%) | 87 (12%) | >0.9 |
| Ethnicity |  |  | 0.076 |
| White British | 109 (72%) | 583 (80%) |  |
| Other | 4 (2.6%) | 13 (1.8%) |  |
| Unknown | 39 (26%) | 136 (19%) |  |
| IMD – median (IQR) | 7 (4, 9) | 6 (4, 9) | 0.4 |
| Unknown | 2 | 9 |  |
| Smoking |  |  | 0.005 |
| Current | 2 (1.3%) | 48 (6.6%) |  |
| Ex-smoker | 84 (55%) | 454 (62%) |  |
| Non-smoker | 54 (36%) | 195 (27%) |  |
| Unknown | 12 (7.9%) | 35 (4.8%) |  |
| Comorbidity scores |  |  |  |
| Rockwood Frailty scale |  |  | 0.13 |
| 1-4 | 45 (38%) | 269 (46%) |  |
| 5-9 | 73 (62%) | 318 (54%) |  |
| Unknown | 34 | 145 |  |
| CCI – median (IQR) | 5 (4, 6) | 5 (4, 6) | >0.9 |
| Respiratory |  |  |  |
| Any^ | 59 (39%) | 322 (44%) | 0.3 |
| COPD | 37 (24%) | 219 (30%) | 0.2 |
| Asthma | 18 (12%) | 89 (12%) | >0.9 |
| Other⁑ | 9 (5.9%) | 73 (10.0%) | 0.13 |
| Cardiovascular |  |  |  |
| Any | 85 (56%) | 408 (56%) | >0.9 |
| IHD | 30 (20%) | 111 (15%) | 0.2 |
| AF | 49 (32%) | 240 (33%) | >0.9 |
| CCF | 31 (20%) | 179 (24%) | 0.3 |
| Diabetes |  |  |  |
| Any | 26 (17%) | 139 (19%) | 0.6 |
| Type 1 | 0 (0%) | 1 (0.7%) | >0.9 |
| Type 2 | 26 (100%) | 138 (99%) |  |
| Neurological |  |  |  |
| Dementia | 26 (17%) | 81 (11%) | 0.041 |
| Cognitive impairment | 18 (12%) | 56 (7.7%) | 0.11 |
| CVA | 20 (13%) | 65 (8.9%) | 0.13 |
| TIA | 18 (12%) | 64 (8.7%) | 0.2 |
| Other neurological disease† | 11 (7.2%) | 34 (4.6%) | 0.2 |
| Immunodeficiency |  |  |  |
| CTD | 16 (11%) | 63 (8.6%) | 0.4 |
| HIV | 0 (0%) | 1 (0.1%) | >0.9 |
| Other immunodeficiency | 20 (13%) | 103 (14%) | 0.9 |
| Oncology |  |  |  |
| Solid organ cancer | 12 (7.9%) | 65 (8.9%) | 0.9 |
| Haematological malignancy | 4 (2.6%) | 21 (2.9%) | >0.9 |
| Renal disease‡ |  |  | 0.5 |
| None | 88 (58%) | 386 (53%) |  |
| Mild | 59 (39%) | 312 (43%) |  |
| Moderate/severe | 5 (3.3%) | 34 (4.6%) |  |

Data are N (%) unless otherwise stated.

*Vaccinated individuals who have received a 5^th^ dose of BA.1/ancestral mRNA bivalent mRNA vaccine after 07/09/22 and less than 3 months prior to admission or a 4^th^ dose of monovalent mRNA vaccine between 21/03/22 and 07/08/22.

#Refers to vaccine brand of the last dose received (5^th^ or 4^th^ dose). Prior to last dose, any vaccine combination is considered.

^Includes any chronic respiratory condition on admission to hospital, such as COPD, asthma, bronchiectasis, pulmonary fibrosis or a rare lung disease
⁑Includes bronchiectasis, pulmonary fibrosis, and other chronic respiratory conditions.
†Includes Parkinson’s disease, Huntingdon’s disease, and other chronic neurological conditions.
‡Mild is CKD stage 1–3; moderate or severe is CKD stage 4–5, end-stage renal failure, or dialysis dependence.

AF, atrial fibrillation; CCF, congestive cardiac failure; CCI, Charlson Comorbidity Index; COPD, chronic obstructive pulmonary disease; CKD; chronic kidney disease; CVA, cerebrovascular accident; IHD, ischaemic heart disease; IMD, Index of multiple deprivation; IQR, interquartile range; LTCF, long-term care facility; TIA, transient ischaemic attack

**Table 3: Relative vaccine effectiveness of 4^th^ dose mRNA monovalent vaccines against hospitalisation, compared to 3^rd^ dose monovalent mRNA vaccines [04/04/22-30/07/22]**

| **Characteristic** | **rVE (95% CI)** | **rOR (95% CI)** | ***P*-value** |
| --- | --- | --- | --- |
| **Univariable Logistic Regression Model** | | | |
| 4th dose of monovalent mRNA vaccine | 51.2 (32.1, 65.0) | 0.488 (0.350, 0.679) | <0.001 |
| **Multivariable Logistic Regression Model** | | | |
| 4th dose of monovalent mRNA vaccine | 46.6 ( 13.9, 67.1 ) | 0.534 ( 0.329, 0.861 ) | 0.011 |
| Time between 3rd dose and admission |  | 1.004 ( 0.999, 1.009 ) | 0.13 |
| Vaccine brand* |  | 0.980 ( 0.636, 1.509 ) | >0.9 |
| Age |  | 1.010 ( 0.981, 1.039 ) | 0.5 |
| Sex (Male) |  | 1.139 ( 0.803, 1.615 ) | 0.5 |
| CCI |  | 0.972 ( 0.873, 1.077 ) | 0.6 |
| IMD |  | 0.974 ( 0.914, 1.037 ) | 0.4 |
| LTCF Resident |  | 0.443 ( 0.221, 0.819 ) | 0.014 |
| Respiratory disease |  | 0.640 ( 0.440, 0.926 ) | 0.019 |
| Prevalence**⁑** |  | 1.001 ( 1.001, 1.002 ) | <0.001 |
| **Matched Conditional Logistic Regression Model†** | | | |
| 4th dose of monovalent mRNA vaccine | 52.0 ( 20.9, 70.9 ) | 0.480 ( 0.291, 0.791 ) | 0.004 |
| Time between 3rd dose and admission |  | 1.005 ( 0.999, 1.010 ) | 0.088 |
| Vaccine brand* |  | 1.002 ( 0.634, 1.584 ) | >0.9 |
| Prevalence**⁑** |  | 1.002 ( 1.001, 1.002 ) | <0.001 |

CI, Confidence Interval; CCI, Charlson comorbidity index; IMD, index of multiple deprivation; rOR, relative Odds Ratio; rVE, relative vaccine effectiveness

* Vaccine brand of 4^th^ dose of monovalent mRNA vaccine or 3^rd^ dose of monovalent mRNA vaccine (prior to last dose, any vaccine combination is considered), where 1= Moderna SpikeVax® and 0 = Pfizer Comirnaty®

**⁑**Prevalence was calculated on a daily basis

**†**1:3 Nearest neighbour propensity score matching with replacement (propensity score estimated using logistic regression on age, sex, CCI score, IMD, LTCF residency and respiratory disease), 179 test-positive cases were matched to 537 test-negative controls with no match found for 138 controls.

**Table 4: Relative vaccine effectiveness of 5^th^ dose BA.1/ancestral mRNA bivalent vaccines against hospitalisation, compared to 4^th^ dose monovalent mRNA vaccines [21/09/22 – 23/01/23]**

| **Characteristic** | **rVE (95% CI)** | **rOR (95% CI)** | ***P*-value** |
| --- | --- | --- | --- |
| **Univariable Logistic Regression Model** | | | |
| 5th dose of bivalent mRNA vaccine | 46.2 (21.1, 63.0) | 0.538 (0.370, 0.789) | 0.001 |
| **Multivariable Logistic Regression Model** | | | |
| 5th dose of monovalent mRNA vaccine | 46.7 ( 18.0, 65.1 ) | 0.533 ( 0.349, 0.820 ) | 0.004 |
| Time between 4th dose and admission |  | 1.000 ( 0.997, 1.003 ) | >0.9 |
| Vaccine brand* |  | 0.882 ( 0.611, 1.283 ) | 0.5 |
| Age |  | 1.006 ( 0.974, 1.038 ) | 0.7 |
| Sex (Male) |  | 1.329 ( 0.925, 1.910 ) | 0.12 |
| CCI |  | 1.005 ( 0.895, 1.123 ) | >0.9 |
| IMD |  | 1.024 ( 0.961, 1.092 ) | 0.5 |
| LTCF Resident |  | 0.953 ( 0.525, 1.646 ) | 0.9 |
| Respiratory disease |  | 0.807 ( 0.551, 1.173 ) | 0.3 |
| Prevalence**⁑** |  | 1.006 ( 0.998, 1.013 ) | 0.13 |
| **Matched Conditional Logistic Regression Model†** | | | |
| 5th dose of monovalent mRNA vaccine | 48.8 ( 19.8, 67.3 ) | 0.512 ( 0.327, 0.802 ) | 0.003 |
| Time between 4th dose and admission |  | 1.000 ( 0.997, 1.003 ) | 0.8 |
| Vaccine brand* |  | 0.909 ( 0.620, 1.335 ) | 0.6 |
| Prevalence**⁑** |  | 1.006 ( 0.999, 1.014 ) | 0.10 |

CI, Confidence Interval; CCI, Charlson comorbidity index; IMD, index of multiple deprivation; rOR, relative Odds Ratio; rVE, relative vaccine effectiveness

* Vaccine brand of 5^th^ dose of BA.1/ancestral mRNA bivalent mRNA vaccine or 4^th^ dose of monovalent mRNA vaccine (prior to last dose, any vaccine combination is considered), where 1= Moderna SpikeVax® and 0 = Pfizer Comirnaty®

**⁑**Prevalence was calculated on a daily basis

**†**1:4 Nearest neighbour propensity score matching with replacement (propensity score estimated using logistic regression on age, sex, CCI score, IMD, LTCF residency and respiratory disease), 150 test-positive cases were matched to 600 test-negative controls with no match found for 123 controls.
